# Dual Contraceptive Method Utilization and Its Associated Factors among women of Reproductive Age at ART Clinics in Lideta Sub-City, Addis Ababa, Ethiopia: A cross-sectional study

**DOI:** 10.1101/2025.08.05.25333092

**Authors:** AsinakeWudu Gessese, Yeshitila Abate Tsehay, Shimels Kimemu Aytenfsu

**Author notes:** Corresponding author Asinake Wudu Gessese.

## Abstract

**Background:** Dual protection is an important preventive approach that prevents both unwanted pregnancy and sexually transmitted infections. It allows Human Immunodeficiency Virus-positive women to avoid unintended pregnancy to reduce vertical HIV transmission, as well as morbidity and mortality among mothers and children. Data regarding dual contraceptive utilization are limited. This study aimed to assess the utilization of dual contraceptive methods and associated factors among reproductive-age women at the ART clinic in Lideta Sub-City, Addis Ababa, Ethiopia, in 2025.

**Methods:** A health center-based cross-sectional study design was employed to collect data from 398 study participants selected through a systematic random sampling technique in eight health centers of Lideta Sub-City from March 7 to April 7, 2025. Binary logistic regression was performed to identify factors associated with dual contraceptive utilization. Adjusted odds ratios with 95% confidence intervals and p-values < 0.05 were used to determine the association between the dual contraceptive utilization and the independent variables.

**Results:** The overall magnitude of dual contraceptive method utilization was 43.2% (95% CI: 38.3%, 48%). Receiving counseling by healthcare providers (AOR=6.25, 95% CI: (1.99, 19.58)), having no desire to have a child (AOR=6.4, 95% CI: (3.24, 12.63)), disclosing HIV status (AOR=4.68, 95% CI: (2.60, 8.43)), and having an open discussion with their partners (AOR=4.3, 95% CI: (1.51, 12.26)) were the factors significantly associated with dual contraceptive utilization.

**Conclusions:** Dual contraceptive utilization is low compared to the targets set by the World Health Organization and the Ministry of Health. Counseling by a health care provider, disclosure of HIV status, desire to have a child, and open partner discussion were the factors associated with dual contraceptive utilization. Hence, educating and empowering HIV-positive individuals regarding their reproductive health choices.

## Introduction

Unintended pregnancy, contraction of new strains of HIV, and pediatric HIV have become challenging issues (1). The consistent and correct use of contraception has far-reaching benefits, including the reduction of pregnancy-related morbidity and mortality, termination of pregnancies, improvement of educational opportunities, and empowerment of women (2, 3).

In Sub-Saharan Africa, increasing the contraceptive prevalence rate (CPR) has been estimated to reduce the proportion of infants infected with HIV by 35–55% through reduction in primary HIV infection and unintended pregnancies in HIV infected women (4). Reports from the United Nations show that contraceptive use has increased globally, as 64% of almost all married women in the regions of the world use a method, compared with Africa, which has 33% and Nigeria at less than 20% practice rate (5).

The World Health Organization (WHO) recommends that women living with HIV use dual protection (6, 7). No single type of contraceptive method is effective at preventing both unintended pregnancies and STIs/HIV at the same time. Thus, dual-contraceptive utilization is the most effective strategy to prevent both unwanted pregnancies and STIs/HIV simultaneously (4, 8, 9). Dual contraceptive utilization is the utilization of barrier contraceptives (i.e., Condoms), associated with another modern contraceptive method (oral contraceptive pills, Jadelle, Depo Provera, Intra-uterine device) (10, 11).

One of the pillars of the World Health Organization (WHO) global effort to prevent Mother to Child Transmission of HIV (MTCT) was the prevention of unwanted pregnancy in HIV infected women (12, 13). To avoid unintended pregnancy and vertical transmission of HIV from mothers to children, dual contraceptive utilization was taken as a better strategy (10).

Dual protection is recommended for all clients on ART to ensure effective and appropriate contraception, as it can prevent both unwanted pregnancy and sexually transmitted infections/HIV (14, 15). Additionally, it can reduce vertical HIV transmission and morbidity and mortality among mothers and children (1).

In 2019, the modern contraceptive prevalence rate of women globally was 91.3% and the low rates were observed in low and middle-income countries (LMICs) as compared to the developed countries (16). Worldwide, HIV is the leading cause of death for women of childbearing age, and up to 64% of all pregnancies are unintended (17). Studies showed that more than 2 million women living with HIV become pregnant every year due to low dual contraceptive method utilization and unsafe sexual practices (1, 10).

Around 90% of HIV infections in children were attributable to Mother to Child Transmission (MTCT) (18). Human Immunodeficiency Virus continues to have disastrous medical, economic, social, and physical impacts on individuals, their communities, and the nations of the world (10). According to data from the global burden of disease study, 5–25% of pregnancy-related deaths globally were due to HIV/AIDS (19). According to the World Health Organization (WHO) report, 21.3% of new pediatric HIV infections are caused by unintended pregnancy among HIV seropositive women (1). More than half of unintended pregnancies among adolescent mothers ended with unsafe abortion (10, 20).

Even though contraceptives have significant importance in reducing HIV transmission and unwanted pregnancies, their use remains low in sub-Saharan Africa (19). The magnitude of dual contraceptive use among HIV-positive women in Cameroon was 33.3% (15). The median contraceptive prevalence among women of reproductive age in sub-Saharan Africa was 28.8% (19). In Sub-Saharan Africa (SSA), a region where reproductive-age women account for the majority of people living with HIV, unintended pregnancies were estimated to account for 14-58 % of all pregnancies (17). The prevalence of unintended pregnancy among HIV-positive women in Uganda was 41.1% (21).

Utilization of dual contraceptives is low in many developing countries, including Ethiopia (22, 23). Across different studies, dual contraceptive utilization among HIV-positive women of reproductive age ranges from 19% to 59.5% (10, 14, 22). According to 2016 Ethiopian Demographic and Health Survey (EDHS), 8% of pregnancies in Ethiopia were not wanted, and 17% of pregnancies were mistimed (10, 24). The pooled prevalence of dual contraception use in Ethiopia was 34.08% (25). More than 90% of pediatric AIDS cases are caused by vertical transmission from mother to child, which also accounts for the majority of heterosexual transmission in Ethiopia (26). The major consequences of family planning failure are increasing new pediatric HIV infection, orphan children, and unwanted pregnancy among these populations (14).

The magnitude of dual contraceptive methods utilization and its determinant factors have not been well understood in resource-limited areas like Ethiopia, specifically in the study area. There were many factors responsible for dual contraceptive utilization for WLHIV that need to be identified by researchers, and these factors differ from facility to facility.

Despite the recognized benefits of dual contraception in preventing unintended pregnancies and improving adherence to ART among HIV-positive women, its utilization remains suboptimal. A significant knowledge gap exists regarding dual contraceptive utilization and the factors associated with its adoption among this vulnerable population. Furthermore, limited evidence is available on the specific barriers and facilitators influencing women’s decisions to utilize dual contraceptive methods, hindering the development of targeted interventions to promote their uptake. This study aims to address this gap by investigating dual contraceptive utilization and identifying the associated factors among HIV-positive women in Health Centers of Lideta Sub-city, Addis Ababa, Ethiopia, 2025, with the ultimate goal of informing strategies to improve reproductive health outcomes and reduce HIV transmission.

## Materials and methods

### Study area and period

The study was conducted in health centers found in Lideta Sub-city, Addis Ababa, Ethiopia, from March 7 to April 7, 2025. Addis Ababa covers an estimated area of 174.4 square kilometers and has an estimated density of 5,535.8 people per square kilometer (27). Based on the United Nations World Urbanization Prospects, the estimated total population projection of Addis Ababa in 2024 was 5,703,630 (28).

Lideta Sub-city has eight health centers and two Hospitals, namely, Balcha Hospital and Police Hospital. In addition, the sub-city has private hospitals and clinics. Private hospitals are Legar Hospital and Amin General Hospital. Private clinics are Tesfa Kokeb, Abinet Clinic, Atlas Clinic, and Connel Medium Clinic. Health centers in the Lideta Sub-City give preventive and curative services to the population of the catchment area.

### Study design and Population

A health Center-based cross-sectional study design was employed. All reproductive age women who are on ART in Lideta sub-city, Addis Ababa, were the source population, while women of reproductive age selected using a systematic random sampling technique, who were on ART in Lideta sub-city Health Centers, Addis Ababa, Ethiopia, were the study population. A selected reproductive-age woman who was on ART was the study unit. All reproductive-age women who were on ART and available during the data collection period were included in the study. However, Women of reproductive-age on ART who were seriously ill were excluded from the study.

### Sample size determination

#### Sample size for the first objective

Sample size was determined using the single population proportion formula and by considering the proportion of dual contraceptive use as 59.5% from the study done in West Shoa Zone on dual contraceptive and factors associated with them among women of ART users (14), and with the assumption of a 95% confidence interval and 5% margin of error. Then, by adding a non-response rate of 10%, the sample size was 408. The final sample size for this study became 408.

#### Sample size for the second objective

Sample size for the objective was calculated using Epiinfo version 7.2.2.6, and with the assumption of 95%confidence level, power 80%, percent of outcome among exposed and non-exposed group, minimum detectable adjusted odds ratio, and non-response 10% (Table 1).

**Table 1:** Sample size calculation for the second objective for the study of dual contraceptive utilization and associated factors among reproductive age group women on ART in Health Centers of Lideta Sub-city, Addis Ababa, Ethiopia, 2025

| S.N | Variables | % in<br>unexposed | % in<br>exposed | AOR | Rati<br>o | Sample<br>size | 10%<br>NRR | Final sample<br>size | Refer<br>ence |
| --- | --- | --- | --- | --- | --- | --- | --- | --- | --- |
| 1 | Counseled by<br>HCP about dual<br>contraceptive | 2.7 | 10.5 | 6.56 | 1:1 | 190 | 19 | 209 | (29) |
| 2 | Having child | 5.9 | 26 | 3.29 | 1:1 | 288 | 29 | 317 | (6) |
| 3 | Residency | 10.8 | 21.6 | 3.64 | 1:1 | 148 | 15 | 163 | (6) |
| 4 | Marital status | 25.8 | 72.3 | 4.33 | 1:1 | 76 | 8 | 84 | (26) |

After calculating the required sample size for the selected variables, the maximum sample size was considered by computing the sample for both objectives. Accordingly, by comparing the sample size for both objectives, the maximum sample size that was used for this study was 408, from the first objective.

### Sampling technique and sampling procedure

There are eight governmental health centers in the Lideta Sub-city, Addis Ababa, Ethiopia. For this study, all eight health centers were included. The sample size for each health center was proportionally allocated based on the client flow in each health center. First, the total number of reproductive age group women data of each health center reviewed and the data revealed that: Hidase Fire Health Center 30, Dagim Hidase Health Center 74, Tekle Haymanot Health Center 93, Abinet Health Center 64, Woreda 04 Health Center 38, General Jagema Kelo Health Center 88, Lideta Health Center 81, and Beletshachew Health Center 48. The total number of reproductive age group women in the eight health centers was 516. While allocating the sample size for each health center proportionally, the sample size for Hidase Fire Health Center 24, Dagim Hidase Health Center 58, Tekle Haymanot Health Center 73, Abinet Health Center 50, Woreda 04 Health Center 30, General Jagema Kelo Health Center 70, Lideta Health Center 64, and Beletshachew Health Center 38.

During the data collection period, study participants were chosen using a systematic random sampling technique, using the sampling interval (K) based on their ART clinic visit calculated by dividing the study population by the sample size. K=516/407=1.2≈1. Therefore, samples were chosen by adding one to the first sample until the required sample size was achieved in all selected health centers (Figure 1).

**Figure 1:**
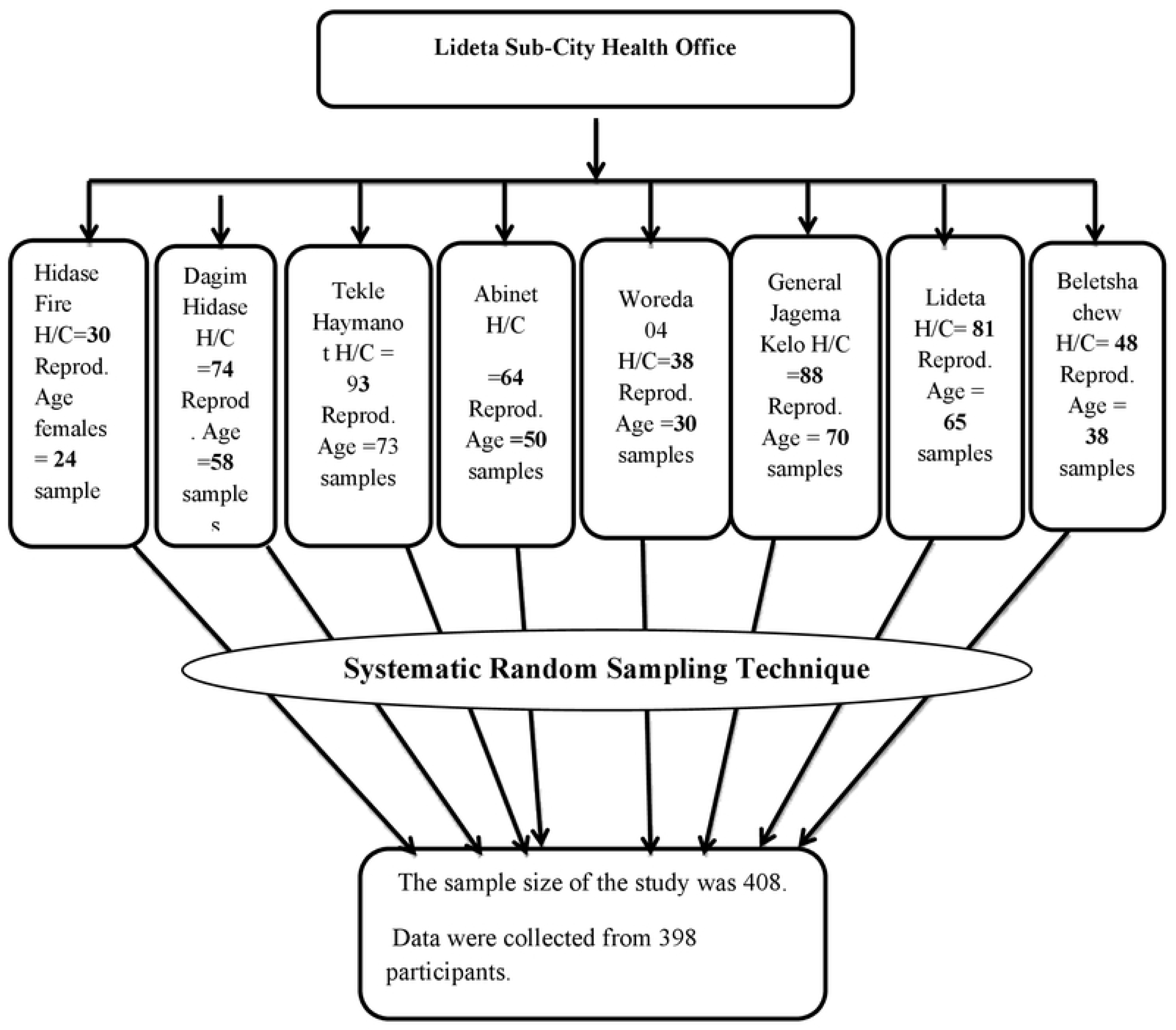
Schematic description of the sampling procedure for the study of dual contraceptive utilization and associated factors among women of reproductive age at the ART clinic in Lideta Sub-city, Addis Ababa, Ethiopia, 2025

### Study variable

#### Dependent variable

Dual contraceptive utilization

#### Independent variables

##### Socio-demographic factors

Age, marital status, educational level, residence, occupation, average monthly income

##### Sexual and reproductive-related variables

Desire of having a child, having a child, freely discussion, post-diagnosis counseling on family planning, family support, sexual active, multiple sexual partners, partner involvement in counseling, STI, counseling by healthcare provider, duration of stay with partner, discussed with t partier their about dual contraceptive utilization, discussed with partner their about dual contraceptive utilization, informed their husband/ partner method of family planning used, discussed with health care providers

##### HIV-related variables

Disclose HIV status to their family, partner starting HAART, partner’s HIV status

#### Operational definition

##### Dual Contraceptive

Utilization of any hormonal or permanent modern contraceptive method along with male or female condoms (30).

##### Current use of the dual method

Sexually active respondents utilizing reversible or irreversible methods of contraception along with male or female condoms during sexual intercourse. **Disclosure of the HIV status:** Disclosing HIV-positive status to the partner and other relatives.

### Data collection tools and procedures

A pretested, close-ended, interviewer-administered structured questionnaire was used for data collection. The questionnaire for the study is adapted based on the literature (10, 22). The questionnaire was designed to address socio-demographic-related factors, sexual and reproductive-related variables, and HIV-related variables. It was translated from English to the local language, Amharic, and back-translated to English to keep internal consistency.

Two days of training were given to orient data collectors and supervisors about the objectives, relevance of the study, confidentiality, the respondents’ rights, informed consent, and interview techniques. Proper supervision was made on data consistency and completeness throughout the data collection process. Data were collected by three trained data collectors and two supervisors. The study populations were invited to participate voluntarily by explaining the rationale of the study at the time of data collection. Trained data collectors and supervisors were used to ask questions and a pretested questionnaire was used for the participants.

#### Data quality assurance

##### Before data collection

The English questionnaire was translated into the local language, Amharic, and then back into English to ensure consistency. Before data collection, the questionnaire was pretested on 5% of the total sample at Ras Emiru Health Center, Arada Sub-City, to determine the response rate, clarity, sequence, and consistency of the questionnaire. The sequence of the tool was adjusted based on the results of the pretest. The data collectors were given two days of training on the study’s objective, relevance, and confidentiality of information, respondents’ rights, informed consent, and interview techniques. In addition, a practical interview demonstration was held in a classroom.

**During data collection,** the data collection and interviewing mechanism was strictly supervised throughout the data collection period by the assigned supervisors and the principal investigator. The questionnaires were checked for completeness and consistency at the site of the data collection by the principal investigator.

**During data entry and analysis,** the collected data were coded and entered into EpiData software version 3.1. The quality of the data was controlled through “skipping patterns”, “must enter” and reduce transportation errors in EpiData. Finally, cleaning and analysis were performed using SPSS version 25.

### Data management and analysis

The collected data were checked for completeness, coded, and entered into EpiData version 3.1, and then exported and analyzed using SPSS computer software package version 25. Data entry was made by the principal investigator. The results of the analysis are presented in the form of text, tables, figures, and summary statistics. For categorical variables, frequencies, percentages, and figures were used. To identify the factors that were associated with dual contraceptive utilization, logistic regression was carried out. Variables with a p-value < 0.25 in the bivariate analysis were candidates for multivariate analysis to control the effect of confounders. Multivariate analysis with a p value < 0.05 was used to estimate associations between dependent and independent variables.

### Ethical consideration

Ethical approval was obtained from the Debre Berhan University, Department of Public Health IRB [Ref.No.: IRB 01/19/2017 E.C, Protocol number: IRB-328], Debre Berhan, Ethiopia, on 5 March 2025. Written informed consent was obtained from each study participant to ensure willingness. Information about the benefits and harms of the study, the usefulness of their participation, the confidentiality of the information, and the right not to participate was given to the participants. The data collectors were given two days of training on the study’s objective, relevance, and confidentiality of information, respondents’ rights, informed consent, and interview techniques in order to address potential ethical vulnerabilities of patients not to feel obliged to participate in the study.

## Results

### Socio-demographic characteristics of respondents

A total of 398 study participants were involved, making a response rate of 97.5%. The mean age of study participants was 29.48 (± 5.9 SD) years, with a minimum of 21 and a maximum of 41 years. Two hundred thirty-three (58.5%) of the respondents are married. Two hundred thirty (57.8%) of the respondents have college and higher educational levels. The mean family monthly income of the respondents was 3957 (±1507 SD) Ethiopian birr. All of the participants are urban residents (**Table 2**).

**Table 2:**
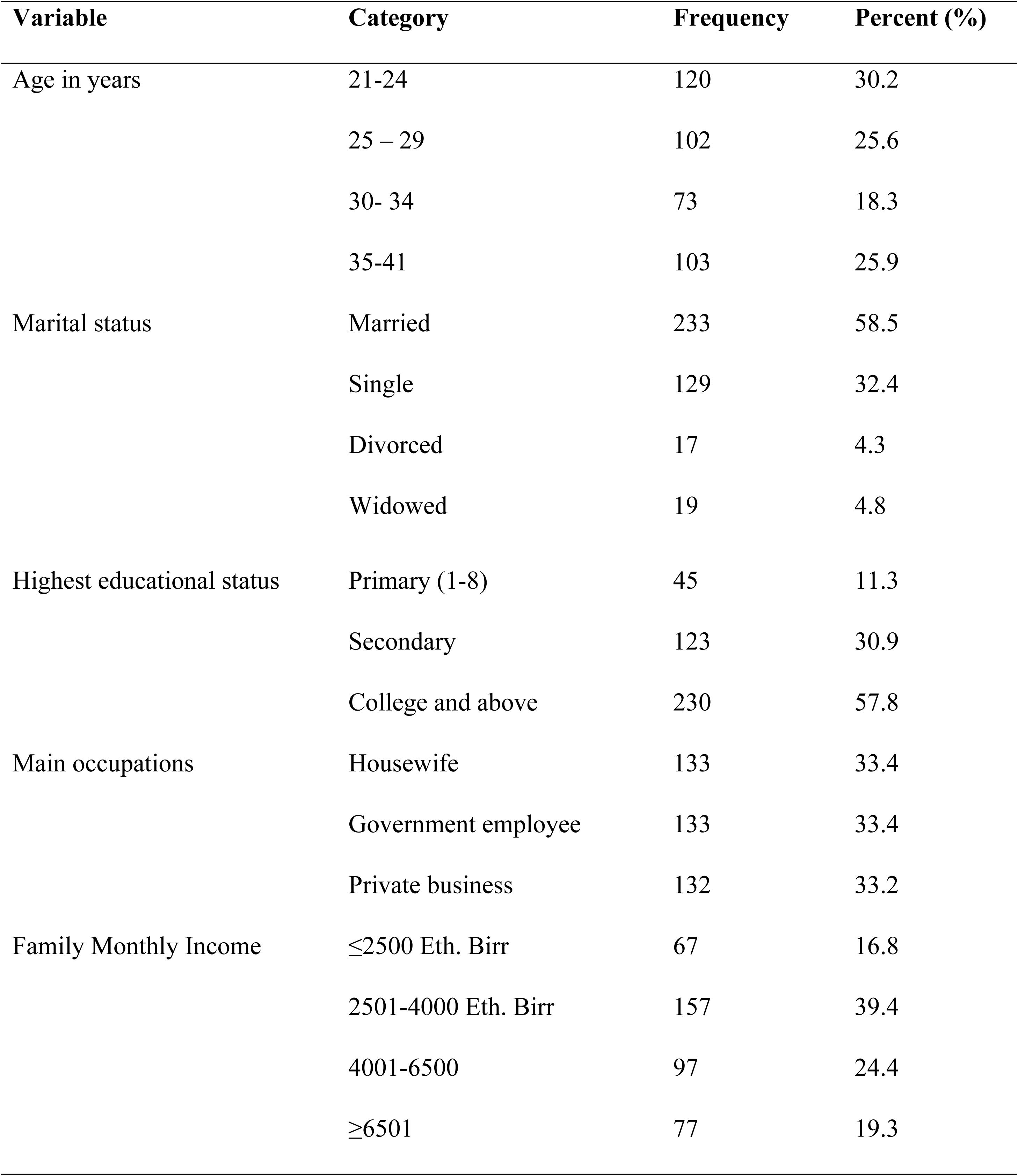
Socio-demographic characteristics of reproductive age group women in ART clinics of Lideta Sub-city Health Centers, Addis Ababa, Ethiopia, 2025 (n=398)

| Variable | Category | Frequency | Percent (%) |
| --- | --- | --- | --- |
| Age in years | 21-24 | 120 | 30.2 |
|  | 25 – 29 | 102 | 25.6 |
|  | 30- 34 | 73 | 18.3 |
|  | 35-41 | 103 | 25.9 |
| Marital status | Married | 233 | 58.5 |
|  | Single | 129 | 32.4 |
|  | Divorced | 17 | 4.3 |
|  | Widowed | 19 | 4.8 |
| Highest educational status | Primary (1-8) | 45 | 11.3 |
|  | Secondary | 123 | 30.9 |
|  | College and above | 230 | 57.8 |
| Main occupations | Housewife | 133 | 33.4 |
|  | Government employee | 133 | 33.4 |
|  | Private business | 132 | 33.2 |
| Family Monthly Income | ≤2500 Eth. Birr | 67 | 16.8 |
|  | 2501-4000 Eth. Birr | 157 | 39.4 |
|  | 4001-6500 | 97 | 24.4 |
|  | ≥6501 | 77 | 19.3 |

### Sexual and reproductive-related variables

Two hundred fifty (62.8%) of the respondents had one or more children. Furthermore, 261 (65.6%) of the participants had a desire to have a child. All the participants are sexually active. Three hundred twenty-five (81.7%) of the respondents had received post-diagnosis counseling about contraceptives. In addition, 266 (66.8%) of the respondents had more than one sexual partner. The mean duration the participants have been with their partner/ husband was 3.58 (±1.96 SD) years (**Table 3**).

**Table 3:** Sexual and reproductive-related variables of reproductive age women in ART clinics of Lideta Sub-city Health Centers, Addis Ababa, Ethiopia, 2025 (n=398)

| Variable | Category | Frequency | Percent (%) |
| --- | --- | --- | --- |
| Do you have a child/ children? | Yes | 250 | 62.8 |
|  | No | 148 | 37.2 |
| Do you have a desire to have a child? | Yes | 233 | 58.5 |
|  | No | 165 | 41.5 |
| Do you have family support? | Yes | 243 | 61.1 |
|  | No | 155 | 38.9 |
| Did you receive post-diagnosis counseling about contraceptives? | Yes | 325 | 81.7 |
|  | No | 73 | 18.3 |
| Have been counseled by healthcare providers about dual contraceptive use | Yes | 253 | 63.6 |
|  | No | 145 | 36.4 |
| Have you discussed with healthcare providers about dual contraceptive use? | Yes | 268 | 67.3 |
|  | No | 130 | 32.7 |
| Have you discussed with your partner/husband about dual contraceptive use? (n=368) | Yes | 119 | 32.3 |
|  | No | 249 | 67.7 |
| Do you have a sexual partner? | Yes | 368 | 92.5 |
|  | No | 30 | 7.5 |
| Was your partner/husband involved in post-test counseling? (n=368) | Yes | 165 | 44.8 |
|  | No | 203 | 55.2 |
| Have you informed your husband/ partner about the family planning method to use? (n=368) | Yes | 130 | 35.3 |
|  | No | 238 | 64.7 |
| Do you have more than one sexual partner? | Yes | 266 | 66.8 |
|  | No | 132 | 33.2 |
| Do you have a history of STIs? | Yes | 260 | 65.3 |
|  | No | 138 | 34.7 |

### HIV-related variables of respondents

Two hundred fifty (62.8%) of the respondents had disclosed their HIV status to their partner and/or family. Additionally, 192 (48.2%) of the respondents’ partners were reactive. The mean duration of ART follow-up of the participants was 7.89 (±2.89 SD) years (**Table 4**).

**Table 4:** HIV-related variables of reproductive age women in ART clinics of Lideta Sub-city Health Centers, Addis Ababa, Ethiopia, 2025 (n=398)

| Variable | Category | Frequency | Percent (%) |
| --- | --- | --- | --- |
| Have you disclosed your HIV status to your partner and/or family? | Yes | 250 | 62.8 |
|  | No | 148 | 37.2 |
| What is your partner's HIV status? (n=368) | Reactive | 176 | 44.2 |
|  | Non-reactive | 119 | 29.9 |
|  | Unknown | 73 | 18.3 |
| If known and reactive, the partner starting HAART (n=176) | Yes | 165 | 41.5 |
|  | No | 11 | 2.8 |

### Dual contraceptive method utilization among women living with HIV attending art clinic in Lideta Sub-City Health Centers

All of the participants had ever heard about family planning. Among those using dual contraceptives, 101 (25.4%) were using condoms with injectables, 64 (16.1%) were using condoms with implants, and 7 (1.8%) of them were using condoms with pills. The overall magnitude of dual contraceptive method utilization was 43.2% (95% CI: 38.3%, 48%) (**Figure 2**).

**Figure 2:**
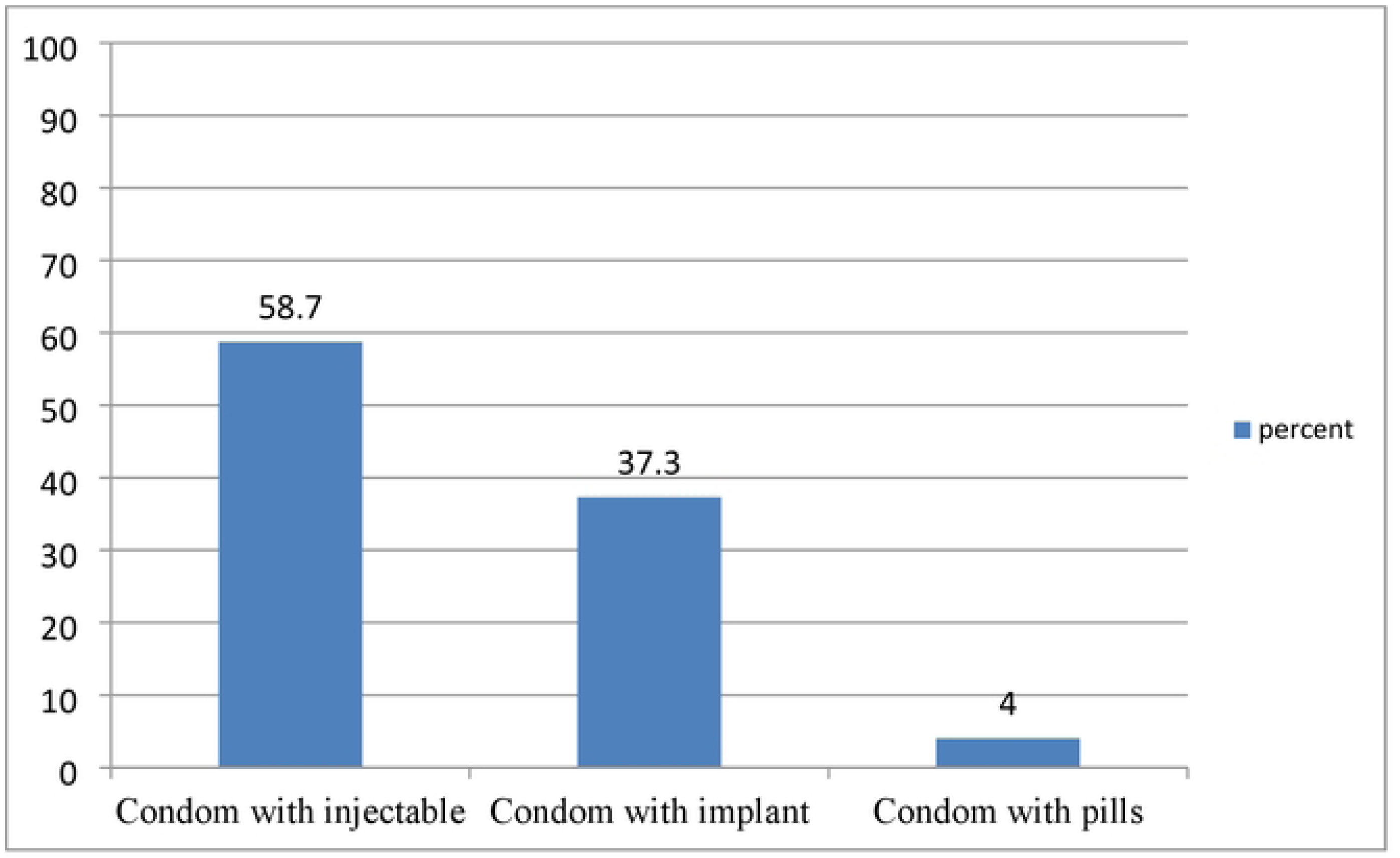
Types of dual contraceptive use among dual contraceptive users in women of reproductive age at ART clinics of Lideta Sub-city Health Centers, Addis Ababa, Ethiopia, 2025 (n=172)

### Factors associated with dual contraceptive utilization

After performing multivariable logistic regression analysis, receiving counseling by a health care provider, desire to have a child, disclosing HIV status, and open partner discussion were significantly associated with dual contraceptive utilization. In this study, participants who were counseled by healthcare providers were six times more likely to use dual contraceptives as compared to their counterparts (AOR=6.25, 95% CI: (1.99, 19.58), P-value=0.002). Similarly, participants who had no desire to have a child/children were six times more likely to use dual contraceptives as compared to their counterparts (AOR=6.4, 95% CI: (3.24, 12.63), P-value=0.000).

Those participants who disclosed their HIV status to friends/family were 4.68 times more likely to use dual contraceptives as compared to their counterparts (AOR=4.68, 95% CI: (2.60, 8.43), p-value=0.000). Finally, those participants who had open discussions with their partners were four times more likely to have dual contraceptive utilization as compared to their counterparts (AOR=4.3, 95% CI: (1.51, 12.26), p-value=0.06) (**Table 5**).

**Table 5:**
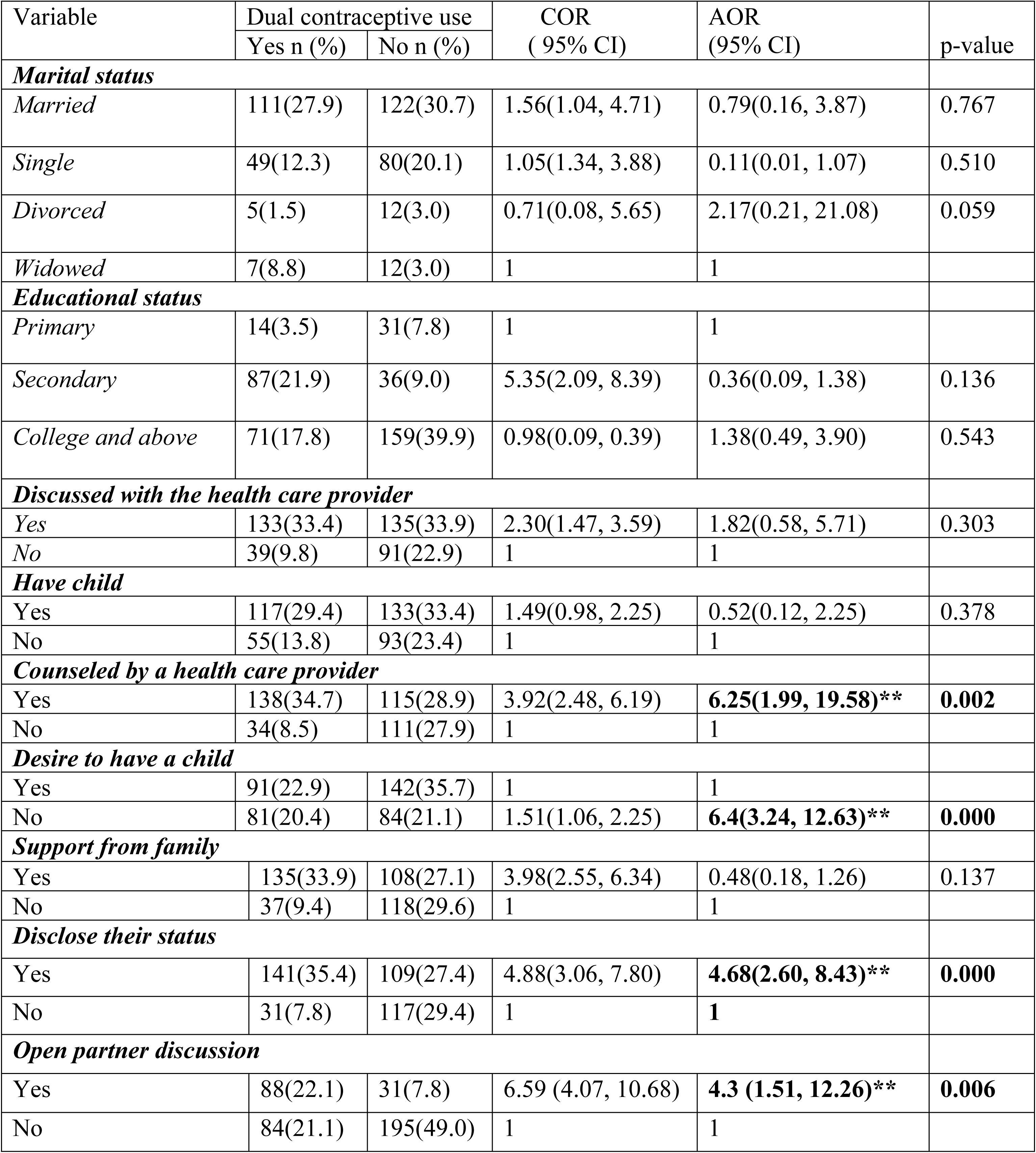
Factors associated with dual contraceptive utilization among reproductive age group women in ART clinics of Lideta Sub-city Health Centers, Addis Ababa, Ethiopia, 2025 (n=398)

## Discussion

The study showed that 43.2% (95% CI: 38.3%, 48%) of the study participants had utilized dual contraceptive methods. Counseling by a health care provider, disclosure of HIV status, desire to have a child, and open partner discussion were the factors associated with dual contraceptive utilization.

This magnitude is in line with a study done in the Eastern Province of Rwanda (40%) (16). The magnitude in this study is higher than a study done in Borena district public health facilities (19.4%) (22), Wolaita Zone (28.6%) (30), Finote Selam Hospital (21.8%) (6), University of Gondar Hospital (13.2%) (29), in North Shewa Zone health facilities (38.1%) (1), and in the West Shewa Zone (21.4%) (10). This discrepancy could be due to the study area between Addis Ababa and the regions, which could have contributed to access to information. In addition, it could be due to socio-demographic and cultural differences of the study populations, the presence of quality, integrated sexual and reproductive health and ART services in Addis Ababa and the regions. Moreover, the difference might be due to the study period. The magnitude in this study is lower than a study done in West Shoa zone Hospitals (59.5%) (14). This discrepancy could be due to the differences in study period and sample size.

Participants who were counseled by health care providers were six times more likely to use dual contraceptives as compared to their counterparts. This finding is supported by studies conducted in the University of Gondar Hospital (29) and in the West Shewa Zone (10). The possible reason could be that healthcare providers offer education about the importance of using dual contraceptives to prevent both HIV transmission and unintended pregnancies. Additionally, counseling provision empowers women to make informed decisions potentially increasing the uptake of dual methods. Counseling also offers information regarding the importance of safer sex, building self-efficacy, and negotiation skills on consistent and correct use of condoms. On the other hand, counseling and providing reproductive health interventions may have a positive influence on minimizing unmet need for dual contraceptive utilization.

Those participants who had no desire to have child/children were six times more likely to use dual contraceptives as compared to their counterparts. This finding is supported by studies conducted in (10, 16, 22, 30). The possible reason could be that dual contraception provides a higher level of protection against unintended pregnancy than a single method. In addition, no contraceptive method is 100% effective. Combining two methods (e.g., condoms and a hormonal method like a pill or implant) significantly reduces the risk of pregnancy compared to using just one.

Those participants who disclosed their HIV status to friends/family were 4.68 times more likely to use dual contraceptives as compared to their counterparts. This finding is supported by studies conducted in North Shewa Zone health facilities (1), West Shewa Zone (10), and Wolaita Zone (30). The possible reason could be that disclosure can facilitate better communication with sexual partners about health status and contraception needs. Additionally, disclosing one’s HIV status can be an empowering act that leads to greater control over one’s sexual health decisions. Moreover, this might be because stigma/discrimination and misconception-related HIV/AIDS were reduced due to community awareness and higher partner care, which leads HIV positive women to disclose their status to their partner. Finally, disclosing HIV status also helps the women to have a common understanding the importance of dual contraceptive utilization and helps to reach an agreement to utilize it.

Those participants who had open discussions with their partners were 4.3 times more likely to have dual contraceptive utilization as compared to their counterparts. This finding is supported by studies conducted in the University of Gondar Hospital (29), in the West Shewa Zone (10), in Hossana (31), and in the Wolaita Zone (30). The possible reason could be that open dialogue fosters better communication about sexual health, desires, and concerns. In addition, when partners openly discuss their health statuses, they can engage in shared decision-making regarding their sexual health. It also encourages HIV status disclosure; future fertility desires, and more importantly, helps to manage life easily. Moreover, open discussion may increase women’s freedom to negotiate with their male partner in the decision of limiting or spacing the number of births and safer sex practices. In conclusion, creating an open discussion between partners about contraceptive choice and its benefits is advantageous.

## Limitations of the study

This study has several limitations. As the study was based on a self-reported data, social desirability bias might be present. Recall bias could also be a limitation since the tool used a self-reported interview. Recall bias could also be present as the tool was self-reported, and it was dependent on their memory; some recall bias might happen. As the study used a cross-sectional study design, it is difficult to relate the temporal relationship. It also has a short duration of study.

## Conclusions

In this study, dual contraceptive utilization is low compared to the targets set by the WHO and the Ministry of Health. Counseling by a health care provider, disclosure of HIV status, desire to have a child, and open partner discussion were the factors associated with dual contraceptive utilization. Healthcare professionals should play a role in educating and empowering HIV-positive individuals regarding dual contraceptive utilization. Open discussions of HIV-positive clients with partners about HIV status and contraceptive use are vital for promoting dual contraceptive utilization. Effective communication fosters trust and understanding, which can lead to more responsible sexual practices among couples.

The healthcare systems should prioritize training for providers on effective counseling techniques that promote open dialogue about sexual health and contraceptive options. Additionally, they should create easily accessible and understandable patient education materials (brochures, videos, etc.) explaining the benefits and risks of various dual contraceptive methods. These materials should be culturally appropriate and address potential barriers to use. Moreover, they should develop strategies to actively involve partners in family planning discussions and decision-making. Finally, implementing peer support programs where women who successfully use dual methods can share their experiences and provide encouragement to others.

Further research is needed to explore the underlying mechanisms that facilitate these relationships. Qualitative studies that provide deeper insights into the personal experiences of HIV-positive individuals regarding counseling, partner communication, and disclosure are recommended.

## Data Availability

All relevant data are within the manuscript and its Supporting Information files.

## Acknowledgments

We would like to forward our deepest gratitude to Debre Berhan University for the chance it has given us to conduct this study. On top of this, we would like to thank the Lideta Sub-city Health Office for giving the background information on the sub-city. Finally, we would like to thank all the data collectors, supervisors and study participants.

## Acronyms and abbreviations

AIDS: Acquired Immune Deficiency Syndrome
ART: Antiretroviral Therapy
CPR: Contraceptive Prevalence Rate
EDHS: Ethiopian Demographic and Health Survey
FSW: Female Sex Workers
HAART: Highly Active Antiretroviral Therapy
HIV: Human Immunodeficiency Virus
PLWHIV: People living with Human Immune Deficiency Virus
STI: Sexually Transmitted Infection
WHO: World Health Organization
WLHIV: Women living with Human Immune Deficiency Virus

## References

1. Debela SM, Adinew YM, Geleta LA, Guye AH. Dual Contraceptive Utilization and Associated Factors among Women Attending Antiretroviral Therapy Clinics in Central Ethiopia, 2022: The Need for a Better Control of Ever-Increasing New Strain of HIV Infection and its Transmission. International Journal of Women’s Health Care. 2023;8(1):39–49.

2. Hlongwa M, Mutambo C, Hlongwana K. ’In fact, that’s when I stopped using contraception’: a qualitative study exploring women’s experiences of using contraceptive methods in KwaZulu-Natal, South Africa. BMJ open. 2023;13(4):e063034.

3. Toguem M. Determinants of Modern Contraception use Among Reproductive Age Women in Cameroon2021.

4. Abay F, Yeshita HY, Mekonnen FA, Sisay M. Dual contraception method utilization and associated factors among sexually active women on antiretroviral therapy in Gondar City, northwest, Ethiopia: a cross sectional study. BMC women’s health. 2020;20(1):26.

5. Ochala E, Syed, Jan SA, Mat S. A LITERATURE REVIEW ON CONTRACEPTIVE PRACTICES, BARRIERS AND MEASURES TO IMPROVE USE AMONG POSTPARTUM WOMEN. 2021.

6. Jemberie A, Aynalem BY, Zeleke LB, Alemu AA, Tiruye TY. Dual Contraceptive Method Utilization and Associated Factors Among HIV Positive Women Attending ART Clinic in Finote-Selam Hospital: Cross-Sectional Study. Archives of Sexual Behavior. 2023;52(6):2639–46.

7. Goshu MT, Haji Y. Dual contraception method use and determinant factors among HIV-positive women of reproductive age in Hawassa, Sidama, Ethiopia, an institutional-based cross-sectional study. Contraception and reproductive medicine. 2025;10(1):36.

8. Yirsaw B, Gebremeskel F, Gebremichael G, Shitemaw T. Determinants of long acting contraceptive utilization among HIV positive reproductive age women attending care at art clinics of public health facilities in Arba Minch town, Southern Ethiopia, 2019: a case control study. 2020;17(1):34.

9. Lee JK, Gutin SA, Getahun M, Okiring J, Neilands TB, Akullian A, et al. Condom, modern contraceptive, and dual method use are associated with HIV status and relationship concurrency in a context of high mobility: A cross-sectional study of women of reproductive age in rural Kenya and Uganda, 2016. Contraception. 2023;117:13–21.

10. Tilahun Y, Bala ET, Bulto GA, Roga EY. Dual contraceptive utilization and associated factors among reproductive-age women on anti-retroviral therapy at hospitals in central Ethiopia. Risk Management and Healthcare Policy. 2021:619–27.

11. Ayele AD, Kassa BG, Beyene FY, Sewyew DA, Mihretie GN. Dual contraceptive utilization and determinant factors among HIV positive women in Ethiopia: a systematic review and meta-analysis, 2020. Contraception and reproductive medicine. 2021;6(1):19.

12. Kefeni B, Tesfaye S, Bayisa K, Gemechu E, Wariso F. Determinants of long act reversible contraceptive utilization among HIV positive reproductive age women attending ART clinic in South West Ethiopia. Contraception and reproductive medicine. 2023;8.

13. Sherwood J, Lankiewicz E, Roose-Snyder B, Cooper B, Jones A, Honermann B. The role of contraception in preventing HIV-positive births: global estimates and projections. BMC Public Health. 2021;21(1):536.

14. Demissie DB, Gudisa T. Dual contraceptive use and associated factors among women living with HIV attending art clinics in West Zone Health Facilities Oromia, Ethiopia. EC Gynaecol. 2019;8(4):143–55.

15. Tsafack M, Essiben F, Momo R, Georges Pius KM, Moyo K, Mpoah Y, et al. Dual Contraception Use and Associated Factors among HIV Positive Women Follow-Up at Treatment Center Unit of Yaounde Central Hospital, Cameroon. OALib. 2020;07:6225.

16. Renzaho JN, Rutayisire E. Dual contraceptive use and associated factors among women aged 15-49 years on antiretroviral therapy in Kayonza District, Rwanda: a cross-sectional study. Pan African Medical Journal. 2022;42(1).

17. Jifar M, Handiso T, Mare T, Ibrahim S. Dual contraceptive utilization and associated factors among Human Immunodeficiency Virus (HIV) positive women attending anti retro viral therapy (ART) clinic in Hossana Hospital, Southern Ethiopia. SM Journal of Gynecology and Obstetrics. 2017;3(2):1023.

18. Ijarotimi O, Ijarotimi I, Ubom A, Balogun M, Fawole O. HIV status and contraceptive use among women of reproductive age group attending a secondary health facility in South-West Nigeria. 2022;7:24–39.

19. Amsalu M, Worku K. Contraceptive use and associated factors among women of reproductive age on antiretroviral therapy in Awabel Woreda health centers, Northwest Ethiopia. 2023;11:20503121231190275.

20. Sutton MY, Zhou W, Frazier EL. Unplanned pregnancies and contraceptive use among HIV-positive women in care. PloS one. 2018;13(5):e0197216.

21. Napyo A, Nankabirwa V, Mukunya D, Tumuhamye J, Ndeezi G, Arach AAO, et al. Prevalence and predictors for unintended pregnancy among HIV-infected pregnant women in Lira, Northern Uganda: a cross-sectional study. Scientific Reports. 2020;10.

22. Amare G, Cherie N, Mekonen AM. Dual contraceptive use and associated factors among reproductive age group on antiretroviral therapy in Borena District, Northeast Ethiopia: a cross-sectional study. HIV/AIDS-Research and Palliative Care. 2021:107–14.

23. Mesfin Y, Argaw M, Geze S, Tefera B. Dual Contraceptive Use and Factor Associated with People Living with HIV/AIDS: A Systematic Review and Meta-Analysis. Infectious Diseases in Obstetrics and Gynecology. 2021;2021.

24. Agency CS, Addis Ababa E, , Program TD, ICF, Rockville M, USA. ETHIOPIA Demographic and Health Survey 2016. July 2017.

25. Ayenew A. Women living with HIV and dual contraceptive use in Ethiopia: systematic review and meta-analysis. Contraception and reproductive medicine. 2022;7(1):11.

26. Bedecha DY, Gurmu MA, Gejo NG. Dual contraception method utilization and associated factors among women on anti-retroviral therapy in public facilities of Bishoftu town, Oromia, Ethiopia. PloS one. 2023;18(1):e0280447.

27. Bureau AACAH. 2023.

28. Prospects UWU. Addis Ababa Demographics. 2024.

29. Reta MM, Tessema GA, Shiferaw G. Prevalence of dual contraceptive use and associated factors among HIV positive women at University of Gondar Hospital, Northwest Ethiopia. BMC research notes. 2019;12:1–7.

30. Haile D, Lagebo B. Magnitude of dual contraceptive method utilization and the associated factors among women on antiretroviral treatment in Wolaita zone, Southern Ethiopia. Heliyon. 2022;8(6).

31. Ashore A, Erkalo D, Prakash R. Dual contraceptives and associated predictors in HIV positive women: a case–control study. Reproductive Health. 2022;19(1):168.

